# Characteristics and outcomes of patients admitted to Swedish intensive care units for COVID-19 during the first 60 days of the 2020 pandemic: a registry-based, multicenter, observational study

**DOI:** 10.1101/2020.08.06.20169599

**Authors:** Michelle S Chew, Patrik Blixt, Rasmus Åhman, Lars Engerström, Henrik Andersson, Ritva Kiiski Berggren, Anders Tegnell, Sarah McIntyre

**Affiliations:** Departments of Anaesthesia and Intensive Care, Biomedical and Clinical Sciences Linköping University Hospital, Sweden; Department of Anaesthesia and Intensive Care, Vrinnevi Hospital, Norrköping, Department of Thoracic and Vascular Surgery, Medical and Health Sciences, Linköping University Hospital, Sweden; Department of Anaesthesia, Intensive Care and Perioperative Services, Umeå University Hospital, Umeå, Sweden; Department of Public Health Reporting, Public Health Agency of Sweden, Sweden; Department of Biomedical and Clinical Sciences, Center for Social and Affective Neuroscience, Linköping University, Sweden

## Abstract

**Background:** The mortality of patients admitted to the intensive care unit (ICU) with COVID-19 is unclear due to variable censoring and substantial proportions of undischarged patients at follow-up. Nationwide data have not been previously reported. We studied the outcomes of Swedish patients at 30 days after ICU admission.

**Methods:** We conducted a registry-based cohort study of all adult patients admitted to Swedish ICUs from 6 March-6 May, 2020 with laboratory confirmed COVID-19 disease and complete 30-day follow-up. Data including baseline characteristics, comorbidities, intensive care treatments, organ failures and outcomes were collected. The primary outcome was 30-day all-cause mortality. A multivariable model was used to determine the independent association between potential predictor variables and the primary outcome.

**Results:** A total of 1563 patients were identified. Median ICU length of stay was 12 (5-21) days, and fifteen patients remained in ICU at the time of follow-up. Median age was 61 (52-69), median Simplified Acute Physiology Score III (SAPS III) was 53 (46-59), and 66·8% had at least one comorbidity. Median PaO_2_/FiO_2_ on admission was 97·5 (75·0-140·6) mmHg, 74·7% suffered from moderate to severe acute respiratory distress syndrome (ARDS). The 30-day all-cause mortality was 26·7%. The majority of deaths occurred during ICU admission. Age, male sex (adjusted odds ratio [aOR] 1·5 [1·1-2·1]), SAPS III score (aOR 1·3 [1·2-1·4]), severe ARDS (aOR 3·1 [2·0-4·8], specific COVID-19 pharmacotherapy (aOR 1·4 [1·0-1·9]), and CRRT (aOR 2·2 [1·6-3·0]), were associated with increased mortality. With the exception of chronic lung disease, the presence of comorbidities was not independently associated with mortality.

**Conclusions:** Thirty-day mortality rate in COVID-19 patients admitted to Swedish intensive care units is generally lower than previously reported. Mortality appears to be driven by age, baseline disease severity, the degree of organ failure and ICU treatment, rather than preexisting comorbidities.

**Funding:** Region Östergötland County Council and Linköping University; number 30320008.

**Research in context:** *Evidence before this study:* In previous studies reporting outcomes for COVID-19 patients admitted to intensive care units (ICUs), none reported 30-day mortality rates, many were censored after short observation periods, and most had substantial proportions of undischarged patients at the time of follow-up. Incomplete data may cause bias in reported mortality rates. Further, national data on critically ill patients have not been previously published.

*Added value of this study:* Our study provides complete 30-day follow up in a nationwide population of 1563 unselected patients admitted to intensive care units in Sweden. All but 15 patients had been discharged from ICU at follow-up thus the study also provides an accurate reflection of ICU mortality. We also provide age-stratified mortality rates and information on ICU treatment and outcomes. This cohort also differs from previous studies in so far as directed antiviral therapy for COVID-19 disease was infrequently used. Adjusted risk estimates for the effect of baseline factors, ICU complications and treatment demonstrate that age, the severity of respiratory failure and need for continuous renal replacement therapy were the most important risk factors for death.

*Implications of all the available evidence:* Mortality rates of COVID-19 patients in Swedish ICUs are lower than those previously reported, despite the high incidence of comorbidities, an ageing population and a high proportion of patients with severe ARDS. Directed antiviral pharmacotherapy was given only to a minority of patients suggesting that survival from COVID-19 in ICU is achievable with good supportive care. Our analysis also suggests that unaccounted factors eg. process and organizational, may be important in determining the outcome of critically ill patients with COVID-19. Our results may be of interest since Sweden has a very limited number of ICU beds and has adopted a unique response to the pandemic compared to other countries. Despite limited numbers of ICU beds per capita, Sweden was able to increase its ICU capacity during the first 2 months of the COVID-19 pandemic and provide essential care to the critically ill with encouraging results.

## Background

Since the initial detection of the Severe Acute Respiratory Syndrome coronavirus–2 (SARS CoV-2) in Hubei Province, China over 16,000,000 cases have been confirmed worldwide.^1^ Admission rates to intensive care units (ICUs) vary substantially, ranging from 5% to 26% in studies from China, Italy and the USA.^2-6^ Explanations for the differences between studies include predisposing factors such as age and comorbidities, availability of ICU beds and mechanical ventilation, as well as ICU admission and treatment practices. The availability of testing also affects the denominator of these statistics, thus the true need for intensive care among SARS-CoV-2 positive persons is difficult to estimate. Yet, knowledge of the incidence and outcomes of critical illness is essential for disaster planning and there is a need to accurately document the healthcare burden for future planning of hospital and ICU surge capacity. Current data in critically ill populations are generally limited in sample size^4,11-14^, with the largest report to date consisting of 1591 patients from the Lombardy Region in Italy.^5^ Better and expedient information with minimal missing data and bias are needed to inform the continued care of COVID-19 patients.

The Swedish Intensive Care Registry (SIR) collects data on patients admitted to ICUs, with 100% coverage since 2019. Thus, SIR is well-poised to report on outcomes in the intensive care population. Little is known about the epidemiology of COVID-19 infections in Sweden, which has one of the world’s highest life expectancies and a significant burden of comorbidity. Sweden also has the second lowest number of ICU beds per capita in Europe and has a low ICU-to-acute care bed ratio.^15^ For comparison, in the first quarter of 2020 Sweden had 5·1 ICU beds per 100000 population, compared to 27/100000 in the USA.^16,17^ The COVID-19 pandemic unleashed a coordinated response in Swedish Intensive Care Units with an increase in the number of beds to 1131 at its peak (pers. comm. Anders Tegnell, Public Health Agency of Sweden), which is twice the pre-pandemic level. Coupled with what is widely perceived to be a ‘relaxed’ national pandemic strategy, results for ICU care in Sweden are understandably under scrutiny. The main goal of this study is to describe demographic characteristics, coexisting conditions, treatments and outcomes among critically ill patients with COVID-19 admitted to Swedish ICUs during the first two months of the pandemic. A secondary goal is to identify independent risk factors associated with increased mortality for these patients.

## Methods

This is a registry-based cohort study of all adult patients (≥18 years) admitted to Swedish Intensive Care Units with PCR confirmed SARS-CoV-2 infection and COVID-19 disease from March 6 to May 6, 2020. The study was approved by the Swedish Ethical Review Authority (no. 2020-01884 and 2020-02498). Written, informed consent was not required. Data on ICU patients are routinely and prospectively uploaded to the SIR. All patients and/or their next-of-kin are informed of the registry and given the possibility to opt-out at any time.

Patients registered in SIR and its supplementary database Swedish Intensive Care Influenza and Viral Infections Registry (SIRI) were eligible. The inclusion period of the study was 6 March (first Swedish COVID-19 case admitted to an ICU) to 6 May 2020 and the population was retrieved from SIR using the diagnosis code U07.1. Only patients with complete 30-day follow-up were included. The first ICU admission during this period for each patient was defined as the index admission. Direct transfer to another ICU was regarded as a continuation of the index admission. Only index admissions were included in the analysis. Data regarding baseline characteristics including comorbidities, intensive care treatments and outcomes were extracted. We also documented ICU length of stay and 30-day and ICU- mortalities. The primary outcome was 30-day all-cause mortality.

## Statistical considerations

Categorical variables are given as frequencies and percentages. Continuous variables are described using medians and interquartile ranges. Simple statistical models are used to provide 95% confidence intervals of the individual measured variables, to facilitate comparisons with similar studies internationally, and future meta-analyses. Mann-Whitney U tests, chi-squared or Fisher’s exact tests are used for comparisons of survivors vs non-survivors.

A multivariable model was used to determine the independent association between predictor variables (e.g. age, sex, comorbidities, Simplified Acute Physiology Score III (SAPS III) score, ICU treatments and complications) and the primary outcome. Predictor variables were chosen on the basis of clinical plausibility and previous studies. Logistic regression was used, and the independent effect (adjusted odds ratio) of different predictor variables estimated from the model.

For categorical variables, a minimum of 10 cases in the smallest category was required for inclusion in a univariable model. For quantitative variables, a minimum n = 30 was required. Only variables with <10% missing data (minimum n = 1407) were included in the multivariable model. Given the exploratory nature of the research, we place more emphasis on the size of the effects found rather than on the outcome of significance tests (although these were performed with a critical p < 0·05 for each test).

Since the degree of preparedness may have been limited during the early stages of the pandemic, we planned a sensitivity analysis for patients admitted only within the first month.

## Results

A total of 2129 case episodes were registered in SIR between 6 March and 6 May, 2020. After removal of duplicates and patients without a Swedish personal identification number, we identified 1563 unique patients admitted to intensive care units in Sweden during the first two months of the SARS-CoV-2 pandemic (Fig 1) with complete 30-day follow-up.

**Figure 1.**
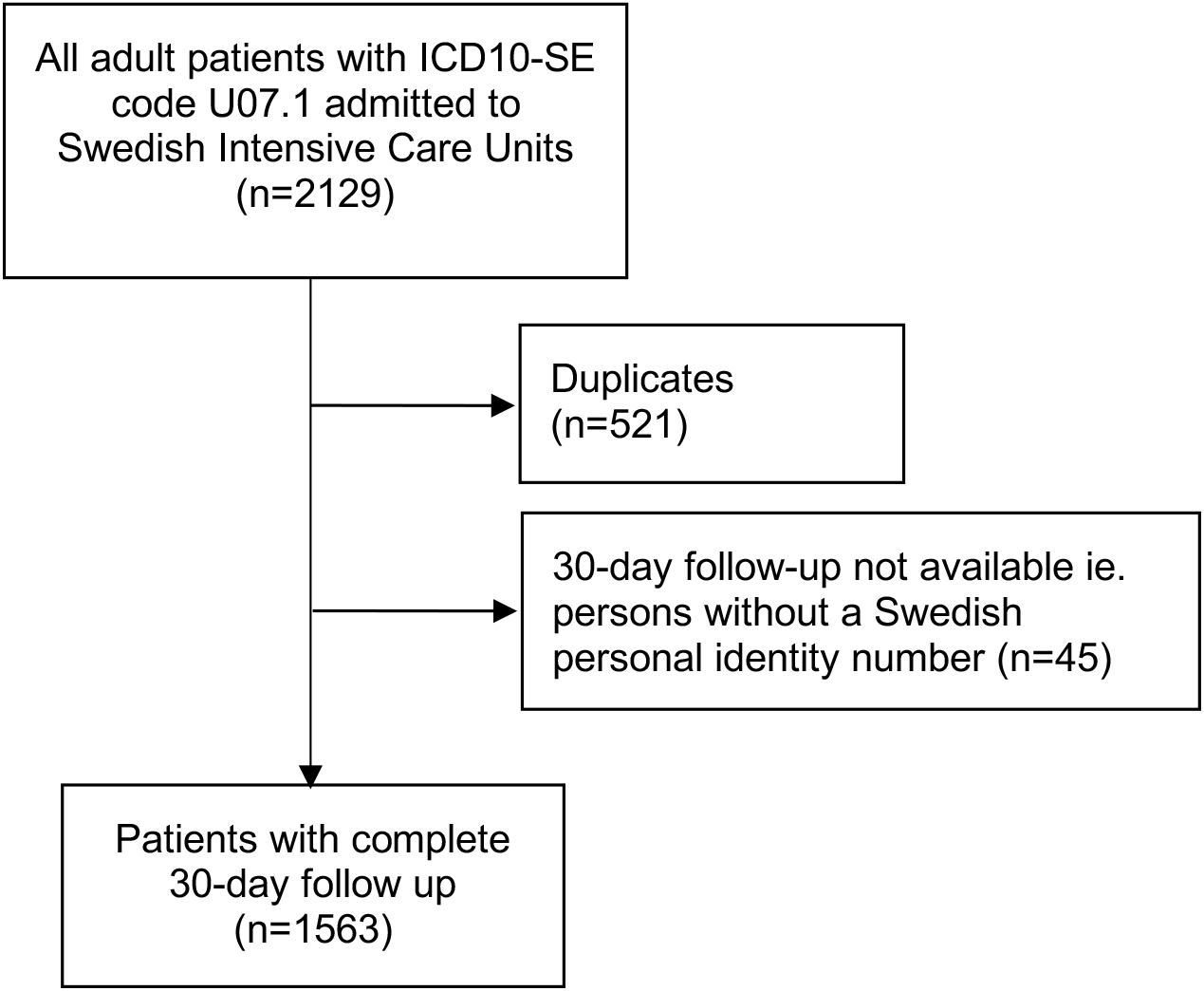
Flowchart for inclusion.

The median age for patients admitted to ICU was 61 (52-69). The median time from symptoms to ICU admission was 10 (7-13) days and the median length of ICU stay was 12 (5- 20) days. A majority has at least one comorbidity (66·8%), with hypertension, diabetes mellitus, obesity (BMI>30) and chronic cardiopulmonary diseases being the most common illnesses. The incidence of Acute Respiratory Distress Syndrome (ARDS) was 80·3%, with 74·7% classified as moderate or severe. Almost half of all patients developed acute kidney injury and 18% received continuous renal replacement therapy (CRRT). ICU and 30-day mortality rates were 23·2% and 26·7% respectively. At 30-day follow up, all but 15 patients had been discharged from ICU, with the majority discharged to a hospital ward. Characteristics and main outcomes of the population are shown in Table 1.

**Table 1.**
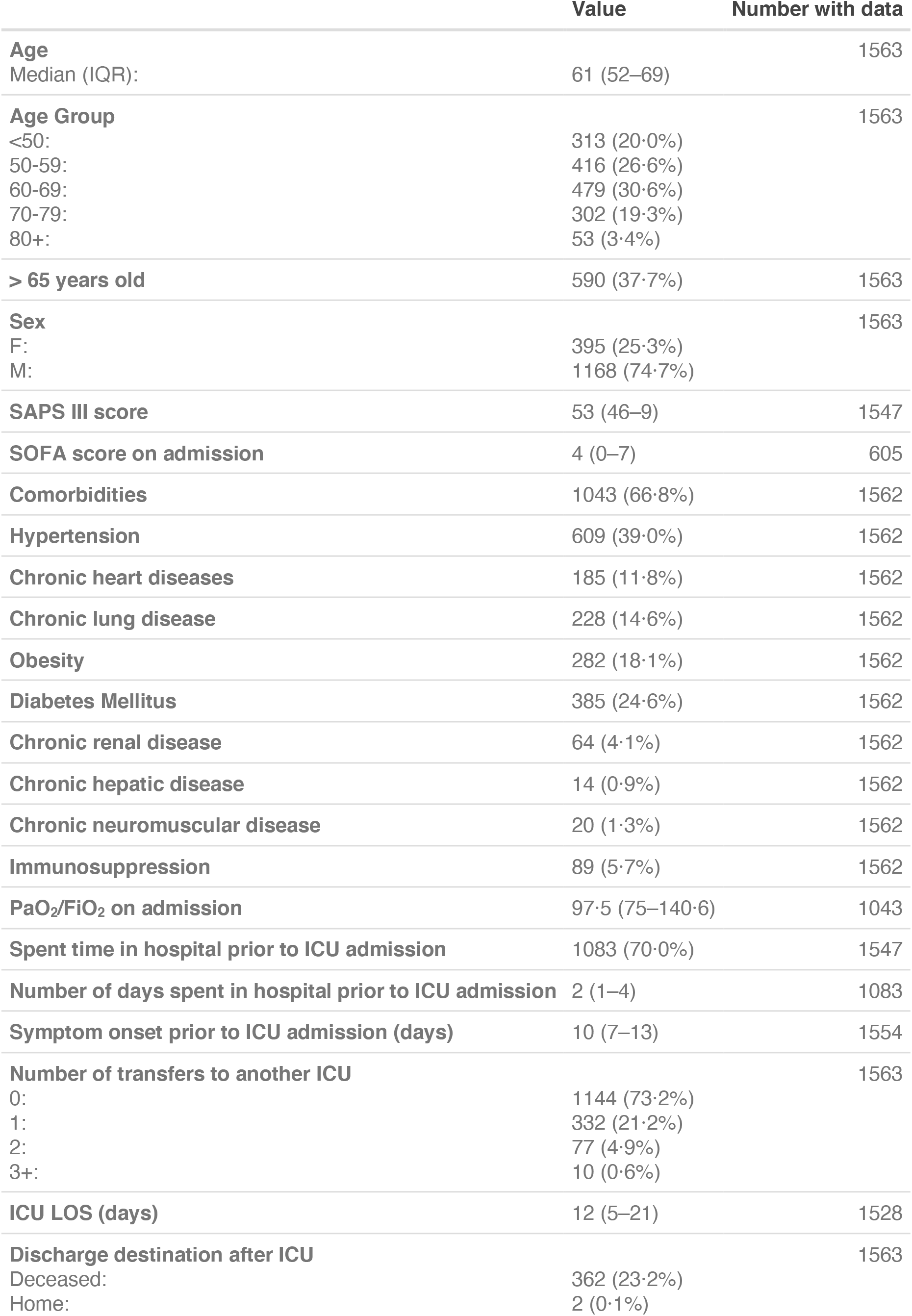
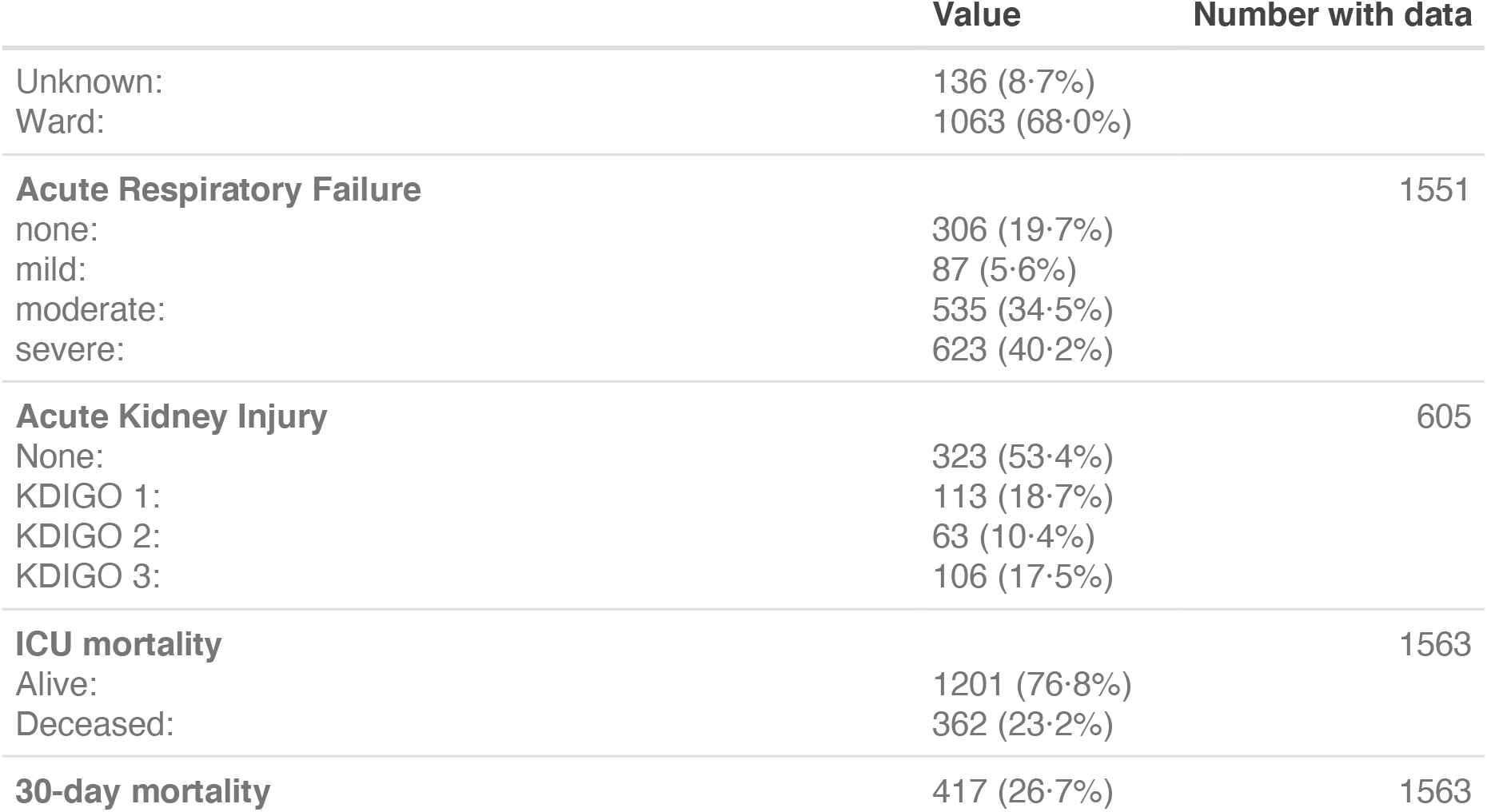
Characteristics of the Swedish COVID-19 ICU population. Data are presented as n (%), or median (IQR).

Treatments given to patients during ICU stay are shown in Table 2. Four-fifths of this cohort received invasive mechanical ventilation, for a median duration of 12 days.

Forty percent of patients were treated with prone positioning, however only a minority received extracorporeal membrane oxygenation. One-fifth of patients received directed treatment for COVID-19 disease, defined as any or more of steroids, remdesivir, tocilizumab, chloroquine/hydroxycholoroquine, lopinavir/ritonavir or ‘other’.

**Table 2.** Treatment received by patients during ICU stay and stratified according to survival. Data are presented as n (%), or median (IQR). P-values indicate differences between survivors and nonsurvivors at 30 days after ICU admission.

| Treatment | N with data |  | Survivors | Non-survivors | p-value |
| --- | --- | --- | --- | --- | --- |
| Invasive mechanical ventilation | 1504 | 1222 (81.2%) | 854/1146 (74.5%) | 368/417 (88.2%) | < 0.0001 |
| No. days of invasive mechanical ventilation | 1222 |  | 13 (8–21) | 11 (5–17) | 0.000228 |
| Noninvasive mechanical ventilation | 1504 | 308 (20.5%) | 240/1146 (20.9%) | 68/417 (16.3%) | 0.0217 |
| High Flow Nasal Oxygen | 1504 | 464 (30.9%) | 377/1146 (32.9%) | 87/417 (20.9%) | < 0.0001 |
| Prone positioning | 1504 | 603 (40.1%) | 404/1146 (35.3%) | 199/417 (47.7%) | < 0.0001 |
| Extra Corporeal Membrane Oxygenation | 1504 | < 20 (<1%) | .. | .. | .. |
| Specific pharmacotherapy | 1562 | 348 (22.3%) | 234/1146 (20.4%) | 114/417 (27.3%) | 0.00386 |
| Hydroxychloroquine/Chloroquine | 1562 | 310 (19.8%) | 201/1146 (17.5%) | 109/417 (26.1%) | 0.000241 |
| Lopinavir/ritonavir | 1562 | < 20 | .. | .. | .. |
| Remdesivir | 1562 | < 20 | .. | .. | .. |
| Tocilizumab | 1562 | 28 (1.8%) | .. | .. | .. |
| Steroids | 1562 | 27 (1.7%) | .. | .. | .. |
| Other | 1562 | 68 (4.4%) | 52/1146 (4.5%) | 16/417 (3.8%) | 0.674 |
| Vasopressors/inotropes | 440 | 191 (43.4%) | 114/1146 (9.9%) | 77/417 (18.5%) | 0.000256 |
| Continuous Renal Replacement Therapy | 1504 | 271 (18.0%) | 150/1146 (13.1%) | 121/417 (29.0%) | < 0.0001 |

SIR reports reintubations and night-time discharges as quality indicators. The incidences of these were 5·2% and 3·6% respectively.

Thirty-day mortality rates increased with increasing age strata: 13·1%, 16·3%, 28·2%, 46·7% and 60·4% for <50, 50-59, 60-69, 70-79 and >80 age groups respectively (χ^2^(4) = 145, p <0·0001). Compared to patients without the comorbidity, 30-day mortality was significantly higher among those with heart disease (37·3% vs 25·2%, p = 0·0007), hypertension (31·2% vs 23·7%, p = 0.001) and pre-existing lung disease (32·5% vs 25·6%, p = 0·0350), but not diabetes mellitus (27·0% vs 26·5%, p = 0·842). Those with obesity had a significantly lower 30-day mortality rate (21·6% vs 27·7%, p = 0·0372; although this was not significant in the multivariable analysis, see Table 3). Patients who were subjected to transfers to other ICUs did not have a higher 30-day mortality (24·6% vs 27·4%, p = 0.273). Compared to survivors, proportionally more nonsurvivors were treated with invasive mechanical ventilation, prone positioning, specific COVID-19 pharmacotherapy and vasopressors or inotropes, and CRRT (Table 2).

Univariable analysis demonstrated an association between the primary outcome and increasing age, sex, most comorbidities, increasing SAPS III and SOFA scores, invasive mechanical ventilation, prone positioning, the use of CRRT, vasopressors and inotropes, as well as specific COVID-19 pharmacotherapy.

Multivariable analysis was applied to 1474 patients with the full dataset. Increasing age, SAPS III score, severe ARDS, the use of CRRT and specific pharmacotherapy against COVID-19 were identified as independent risk factors for 30-day all-cause mortality. The use of high flow nasal oxygen was protective, an effect retained even after adjustment for confounders. Except for chronic lung disease, none of the other registered comorbidities were associated with the primary outcome after adjustment (Table 3).

**Table 3.** Univariable and multivariable analysis of risk factors for 30-day mortality. * p < 0·05, ** p < 0·01, *** p < 0·001)

| Variable | Univariable OR (95% CI) | Multivariable OR (95% CI) |
| --- | --- | --- |
| **Age Group**<br>(Reference: <50 years) |  |  |
| 50-59 | 1.3 (0.9–2.0) | 1.0 (0.6–1.7) |
| 60-69 | 2.6 (1.8–3.9)*** | 1.9 (1.2–3.0)** |
| 70-79 | 5.8 (3.9–8.7)*** | 3.9 (2.4–6.5)*** |
| 80+ | 10.1 (5.4–19.5)*** | 6.5 (2.9–14.7)*** |
| **Male** | 1.4 (1.1–1.9)** | 1.5 (1.1–2.1)* |
| **SAPS III score**<br>OR per increase of 5 points | 1.5 (1.4–1.6)*** | 1.3 (1.2–1.4)*** |
| **SOFA score**<br>OR per increase of 1 point | 1.2 (1.1–1.2)*** | .. |
| **Obesity** | 0.7 (0.5–1.0)* | 0.9 (0.6–1.3) |
| **Diabetes Mellitus** | 1.0 (0.8–1.3) | 1.0 (0.7–1.4) |
| **Hypertension** | 1.5 (1.2–1.8)** | 1.0 (0.7–1.3) |
| **Chronic heart diseases** | 1.8 (1.3–2.4)*** | 1.0 (0.6–1.5) |
| **Chronic lung disease** | 1.4 (1.0–1.9)* | 1.6 (1.1–2.3)* |
| **Chronic renal disease** | 1.3 (0.7–2.1) | 0.7 (0.3–1.4) |
| **Immunosuppression** | 2.0 (1.3–3.0)** | 1.4 (0.8–2.4) |
| **Time from symptom onset to ICU**<br>OR per increase of 7 days | 0.7 (0.6–0.8)*** | 0.7 (0.5–0.8)*** |
| **PaO<sub>2</sub>/FiO<sub>2</sub> on admission**<br>OR per increase of 37.5 mmHg | 0.9 (0.9–1.0)* | .. |
| **Acute Kidney Injury**<br>(Reference: none) |  |  |
| KDIGO 1 | 0.9 (0.5–1.5) | .. |
| KDIGO 2 | 1.8 (1.0–3.1)* | .. |
| KDIGO 3 | 1.9 (1.2–3.0)** | .. |
| **Acute respiratory failure**<br>(Reference: none) |  |  |
| mild | 0.9 (0.4–1.6) | 1.3 (0.6–2.6) |
| moderate | 1.0 (0.7–1.4) | 1.0 (0.6–1.5) |
| severe | 3.0 (2.2–4.3)*** | 3.1 (2.0–4.8)*** |
| **Invasive mechanical ventilation** | 2.4 (1.7–3.4)*** | 1.2 (0.8–1.9) |
| **Noninvasive mechanical ventilation** | 0.7 (0.5–0.9)* | 0.6 (0.4–0.8)** |
| **High Flow Nasal Oxygen** | 0.5 (0.4–0.7)*** | 0.5 (0.4–0.7)*** |
| **Prone positioning** | 1.6 (1.3–2.0)*** | 1.4 (1.0–1.8)* |
| **Specific pharmacotherapy<sup>1</sup>** | 1.5 (1.1–1.9)** | 1.4 (1.0–1.9)* |
| **Vasopressors/Inotropes** | 2.2 (1.4–3.3)*** | .. |
| Continuous Renal Replacement Therapy | 2.6 (2.0–3.4)*** | 2.2 (1.6–3.0)*** |
| Transfer to other ICU | 0.9 (0.7–1.1) | 0.8 (0.6–1.1) |

<sup>1</sup>any or more of chloroquine/hydroxychloroquine, lopinavir/ritonavir, remdesivir, Tocilizumab, steroids, or ‘other’

A sensitivity analysis limiting the cohort to patients admitted within the first 30 days of the pandemic indicated that 30-day mortality was higher during this period compared to the second 30 days (OR 1·9 [1·5-2·4]), Fisher’s Exact Test, p-value < 0.0001) with no interaction between admission period and the other terms in the multivariable model (Likelihood ratio test, χ^2^ = 32·5, p = 0·115). Risk estimates for all covariates were similar compared to the whole cohort (Supplementary Table 1).

## Discussion

In this study of early admissions to Swedish ICUs, we found a 30-day all-cause mortality of 26·7% and ICU mortality of 23·2% for adult patients admitted with COVID-19. With the exception of a case series from Vancouver, Canada^10^, these mortality rates are generally lower than those previously reported in ICU populations. Comparison of mortality rates across studies is difficult due to different lengths of follow-up and significant proportions of patients remaining in ICU at follow-up in many studies.^5,6,8,11,12,14^ Our results compare favorably to a recent systematic review that reported an ICU mortality of 41·6% for patients with completed ICU admissions.^14^ We also noted a decrease in mortality rates in the second half of our population. Our results add to current knowledge by providing a longer and complete follow-up for an unselected, nationwide ICU population. Characteristics and outcomes of the present population, and those of other ICU populations are summarized in Supplementary Table 2. The 30-day mortality rate is only slightly higher than ICU mortality rates for our population, indicating that most patients succumb during ICU care.

We confirm previous findings^4,5,7,10,13^ that mortality rates are significantly higher among those aged >65 years. Very old patients (≥80 years of age) made up 3·4% of all admissions, within the range reported in previous studies.^4,5,9,13^ The mortality in this group was high, 60·4% in our study compared to 35-100% in Wuhan^4,13^, 63% in New York^9^, and 55% in Lombardy^5^ although the latter is likely an underestimation with 36% of patients still in ICU at the time of data collection. These data should however be interpreted with caution since sample sizes are very small and not likely representative of the population means. Although octogenarians have a >6·5-fold adjusted risk of death compared to those <50 years of age, our data demonstrate that provision of intensive care should not be restricted on the basis of age alone.

A majority of patients suffered from comorbidities, the most commonly hypertension, diabetes and obesity. Whilst most comorbidities were associated with the primary outcome in univariable analyses, their effects were no longer significant after multivariable adjustment. The exception was chronic lung disease, which was associated with a 1·6-fold risk of death. These findings are consistent with a recent case series from New York^9^ and deviate from other studies that have not adjusted for confounders.^4,5^ Obesity was not associated with increased mortality as suggested by other studies.^18,19^ These differences may be due to differences in adjustment for other confounders and risk modifiers, or the definition of obesity.

The median ICU length of stay (LOS) for this cohort exceeds that for ICU patients in Sweden prior to the pandemic.^20^ As a comparison, the median LOS for influenza cases during 2015-19 was 3·65 days.^21^ We also observed a higher incidence of AKI than has been previously reported^13,22^, and have identified only one other study with reported incidences in excess of 30%^11^. Of those with documented AKI, 38% suffered from severe kidney injury defined as Kidney Disease Improving Global Outcomes (KDIGO) stage 3. We can only speculate as to the causes of the high incidence of AKI, which may be due to a direct effect of SARS-CoV-2 on the kidneys or a consequence of treatment in intensive care such as fluid restriction and nephrotoxic agents. We demonstrate for the first time that CRRT is independently associated with 30-day mortality in critically ill patients with COVID-19, with a 2-fold increased adjusted risk. Our findings support those observed by Cheng et al. that demonstrated an independent association between the degree of AKI and in-hospital mortality.^23^ These findings highlight the importance of potentially preventable complications for survival outcomes.

Eighty percent of patients received invasive mechanical ventilation, less than those reported in Lombardy^5^ (88%) and Vitoria^12^ (94%) despite more severe hypoxemia on admission in our population. The high proportion of moderate-severe ARDS in this population may be a consequence of Swedish ICU admission policies. Of note, only severe ARDS was independently associated with mortality. A larger proportion (40%) patients were cared for in the prone position compared to previous reports^4,5,9,10^, possibly reflecting the availability of nursing resources. Swedish ICUs employ nurses’ aides to facilitate nursing care, and during the pandemic medical students were also recruited to assist in this role. The use of high flow nasal oxygen (HFNO) was protective even after adjustment for baseline disease severity and degree of ARDS. However, unaccounted confounders may explain this effect, and our experience is that most patients were intubated expediently either on arrival to, or prior to ICU admission due to the severity of hypoxemia, and not often extubated to HFNO.

Another noteworthy observation was that only 22% of patients received specific pharmacotherapy for COVID-19 disease compared to 38-100% in previous reports (Supplementary Table 3).^4,5,6,7,11-13^ The vast majority were treated with Chloroquine or Hydroxychloroquine during the first month of data collection. Only five patients received this drug in the second month of data collection consistent with compliance with the European Medicines Agency directive published on 1 April, 2020 for use of the Chloroquine and Hydroxychloroquine only within clinical trials of emergency use programs. Although we demonstrated an increased risk of death associated with COVID-19 pharmacotherapy in the whole cohort, this finding should be interpreted cautiously since the present study was not designed to assess such an effect.

We chose a simple, exploratory, multivariable model with covariates chosen based on previous findings, clinical plausibility and the availability of data. In general, 30-day mortality was less driven by preexisting comorbidities than variables reflecting severity of organ failure such as ARDS and CRRT. Unfortunately, the degree of missing data for treatment with vasopressors and inotropes precluded its entry into the multivariable analysis. Nevertheless, the avoidance of further deterioration of pulmonary and renal function seems to be a prudent strategy for supportive care, in the absence of specific COVID-19 therapy. We are encouraged by the generally lower mortality rates than previously reported, that was achievable by supportive care. We hypothesize that process and organizational factors have likely contributed to the relatively good outcomes seen in Swedish ICUs as staffing, protective equipment, availability of drugs, medical and technical equipment were considered within the pandemic plans at an early stage at hospital and regional levels.

The ICU admission rate among test positive persons for the first two months of the pandemic in Sweden was 6·5% (1608 of 24722 test positive on 6 May 2020, pers. comm. Anders Tegnell, Public Health Agency of Sweden) ^24^ similar to that described by the Chinese Centre for Disease Control, where 5% of 72314 confirmed cases of COVID 19 were classified as ‘critical’ ie. respiratory failure, septic shock and/or multiple organ failure.^3^ Other studies document ICU admission rates of 8% among hospitalized patients in a single center in Wuhan^4^, 14·2% among hospitalized patients in New York^2^ and 9% among test positive persons in the Lombardy region^5^. The lower proportion of ICU admissions due to COVID-19 in Sweden may reflect the very limited availability of ICU beds, and organizational differences between countries.

The encouraging outcomes for the intensive care population are in contrast to those reported from nursing and aged care facilities who make up the majority of deaths in SARS-CoV-2 positive patients in Sweden.^25^ We believe it is unlikely that intensive care admission would have been effective in preventing these deaths, given the extremes of frailty, age and comorbidities in this population group. These results should also be interpreted within the wider context of SARS-CoV-2 infections in Sweden. The pandemic response in Sweden, with a population of 10 million, has been different from other countries, with emphasis on individual social responsibility. The main strategy is to maintain hygiene, practice distancing to avoid close contacts, limit the size of social gatherings, as well as voluntary closure of large winter resorts and mandatory closure of some leisure centres, with enforcement of distancing rules in restaurants. The results of this approach appear to have been successful in so far as the health care system capacity in Sweden has not been overwhelmed. The number of available ICU beds was doubled but the proportion of occupied ICU beds in the country during the study period never reached maximal capacity (pers. comm. Anders Tegnell, Public Health Agency of Sweden). To cope with the increased flow of patients, specific COVID-19 disaster management plans were activated and all hospitals repurposed other units to admit COVID-19 patients requiring intensive care. Intermediary care units were opened. A massive educational effort was launched to prepare physicians and nurses not traditionally involved in the care of the critically ill to provide this level of care under the supervision of intensivists. Also, anesthesia and intensive care are combined specialities in Sweden, and physicians have dual competence in these areas which facilitated rapid diversion of resources from perioperative to intensive care. Sweden also has a large number of nurse-anesthestists, who, although not specifically trained in intensive care, were expediently deployed for care of mechanically ventilated COVID-19 patients after short ‘refresher’ courses. Elective surgical procedures were postponed and many postoperative care units were converted to pandemic ICUs instead. This mitigated situations of acute staffing, bed and equipment shortages. While this has been advantageous in the short-term, the longer-term effects are still unknown. Future staffing shortages, the looming possibility of burn-out and numerous organizational challenges remain. A significant burden is the management patients that did not receive care during the pandemic. For example, Sweden now has a massive backlog of elective surgical procedures with a cumulative reduction of 60% during the first 2 months of the study.^26^

Limitations of this study include its retrospective nature, although data was prospectively registered in a large centralized database. We were unable to explore outcomes related to ethnicity or socioeconomic status as these are not registered in SIR. Specific data on laboratory variables such as lymphocyte counts and inflammatory parameters, ventilatory modes, sedation and analgesia and imaging were not recorded. Complications related COVID-19 are reported by each unit but are not validated and the degree of missing data varies widely from 100% for delirium and myocardial injury to 61% for acute kidney injury. Important complications such as venous thromboembolism, cardiac injury and cognitive dysfunction at discharge are not systematically queried and are likely to be substantially underreported in the SIRI database. While we make observations regarding pharmacological treatment and availability of ICU beds, any relationship with outcomes are purely speculative. Finally, we only capture data for the first two months of ICU admission and reported outcomes may change over the course of time; and may not be generalizable outside Sweden.

The strengths of this study are that it encompassed an unselected population in Sweden, and captured the peak of COVID-19 admissions to intensive care units. This is significant in so far as it covers the period during which the intensive care capacity was maximally stressed, with possible consequences of outcome. We noted a significantly higher 30-day mortality during first 30 days of the study period. This was an effect of admission period only, as there were no interactions between this and any of the other covariates, and the risk estimates for individual covariates remained similar throughout. Thus, unaccounted confounders (eg. availability of beds, personnel and familiarity with COVID-19 care, differences in supportive treatments) are likely to be in play. Data beyond 30 days are not yet available and it would be relevant to investigate long-term outcomes to assess the true burden of COVID-19 disease.

All but 15 patients had been discharged from ICU at follow-up thus the study also provides an accurate reflection of ICU mortality. Another strength of this study is the full coverage of SIR during our inclusion period (100% from 2019, pers. comm. Riitva Kiiski Berggren, Swedish Intensive Care Registry), completeness and accuracy of the routinely collected variables and validation of these basic ICU data by SIR. All eligible patients were included thus selection bias is minimal. Mortality outcomes were available for all patients, except 45 persons without a Swedish personal identification number, and verified in the Swedish Registry of Deaths. This is in contrast to previous studies where data were censored at various timepoints after ICU admission and with substantial proportions of patients still in ICU at follow up.^5,9,11,12^

## Conclusion

In conclusion, mortality rates in COVID-19 patients admitted to Swedish intensive care units are generally lower than previously reported. Mortality appears to be driven by age, baseline disease severity, the degree of organ failure and ICU treatment, rather than preexisting comorbidities.

## Data Availability

Our group is committed to open science. The data from this study will be made available after publication, upon application to the corresponding author and within the terms of the Global Data Protection Regulation and the Swedish Patient Data Law (2008:355). To avoid the possibility of identifying individual cases, detailed data are not given in the paper or appendix but may be requested from the corresponding author.

## Contributors

MSC conceived the study. All authors contributed to its methodology and statistical analysis plan. SM conducted all statistical analyses. MSC drafted the initial manuscript and the document was edited and approved by all coauthors.

## Acknowledgments

We are indebted to all contributing hospitals for uploading data to the SIR. We also gratefully acknowledge the efforts of Caroline Mårdh from the Swedish Intensive Care Registry for ensuring the most complete and updated dataset possible, and for data extraction.

